# Age, sex and *APOE*-ε4 modify the balance between soluble and fibrillar β-amyloid in cognitively intact individuals: topographical patterns and replication across two independent cohorts

**DOI:** 10.1101/2020.12.03.20241851

**Authors:** Raffaele Cacciaglia, Gemma Salvadó, José Luis Molinuevo, Mahnaz Shekari, Carles Falcon, Gregory Operto, Marc Suárez-Calvet, Marta Milà-Alomà, Arianna Sala, Elena Rodriguez-Vieitez, Gwendlyn Kollmorgen, Ivonne Suridjan, Kaj Blennow, Henrik Zetterberg, Juan Domingo Gispert, for the Alzheimer’s Disease Neuroimaging Initiative°, for the ALFA study

**Affiliations:** Barcelonaβeta Brain Research Center (BBRC), Pasqual Maragall Foundation, 08005 Barcelona, Spain; Hospital del Mar Medical Research Institute (IMIM), 08005 Barcelona, Spain; Centro de Investigación Biomédica en Red de Fragilidad y Envejecimiento Saludable (CIBERFES), 28089 Madrid, Spain; Universitat Pompeu Fabra, 08002 Barcelona, Spain; Centro de Investigación Biomédica en Red de Bioingeniería, Biomateriales y Nanomedicina (CIBERBBN), 28089 Madrid, Spain; Servei de Neurologia, Hospital del Mar, Barcelona, Spain; Department of Neurobiology, Care Sciences and Society, Division of Clinical Geriatrics, Center for Alzheimer Research, Karolinska Institutet, 141 52 Stockholm, Sweden; Roche Diagnostics GmbH, Penzberg, Germany; Roche Diagnostics International Ltd, Rotkreuz, Switzerland; Department of Psychiatry and Neurochemistry, Institute of Neuroscience and Physiology, The Sahlgrenska Academy at the University of Gothenburg, 41390 Mölndal, Sweden; Clinical Neurochemistry Laboratory, Sahlgrenska University Hospital, 41390 Mölndal, Sweden; UK Dementia Research Institute at UCL, WC1E 6BT London, UK; Department of Neurodegenerative Disease, UCL Institute of Neurology, WC1N 3BG London, UK

## Abstract

Cerebral beta-amyloid (Aβ) accumulation is the earliest detectable pathophysiological event along the Alzheimer’s disease (AD) *continuum*, therefore an accurate quantification of incipient Aβ abnormality is of great importance to identify preclinical AD. Both cerebrospinal fluid (CSF) Aβ concentrations and Position Emission Tomography (PET) with specific tracers provide established biomarkers of Aβ pathology. Yet, they identify two different biological processes reflecting the clearance rate of soluble Aβ as opposed to the cerebral aggregation of insoluble Aβ fibrils. Studies have demonstrated high agreement between CSF and PET-based Aβ measurements on diagnostic and prognostic levels. However, an open question is whether risk factors known to increase AD prevalence may promote an imbalance between these biomarkers, leading to a higher cumulative Aβ cerebral aggregation for a given level of cleared Aβ in the CSF. Unveiling such interactions in cognitively unimpaired (CU) individuals shall provide novel insights into the biological pathways underlying Aβ aggregation in the brain and ultimately improve our knowledge on disease modelling. With this in mind, we assessed the impact of three major unmodifiable AD risk factors (age, *APOE*-ε4 and sex) on the association between soluble and deposited Aβ in a sample of 293 middle-aged CU individuals who underwent both lumbar puncture and PET imaging using the [^18^F]flutemetamol tracer. We looked for interactions between CSF Aβ42/40 concentrations and each of the assessed risk factors, in promoting Aβ PET uptake both in candidate regions of interest and in the whole brain. We found that, for any given level of CSF Aβ42/40, older age and female sex induced higher fibrillary plaque deposition in neocortical areas including the anterior, middle and posterior cingulate cortex. By contrast, the modulatory role of *APOE*-ε4 was uniquely prominent in areas known for being vulnerable to early tau deposition, such as the entorhinal cortex and the hippocampus bilaterally. *Post hoc* three-way interactions additionally proved evidence for a synergistic effect among the risk factors on the spatial topology of Aβ deposition as a function of CSF Aβ4/40 levels. Importantly, findings were replicated in an independent sample of CU individuals derived from the ADNI cohort. Our data clarify the mechanisms underlying the higher AD prevalence associated to those risk factors and suggest that *APOE*-ε4 in particular paves the way for subsequent tau spreading in the medial temporal lobe, thus favoring a spatial co-localization between Aβ and tau and increasing their synergistic interaction along the disease *continuum*.

## INTRODUCTION

Alzheimer’s disease (AD) is characterized by cerebral accumulation of misfolded amyloid beta (Aβ) and tau proteins along with progressive neuronal degeneration. AD has an insidious onset, with a protracted asymptomatic phase lasting about two decades prior to clinical manifestations (Sperling et al., 2011). According to recent pathophysiological models, Aβ pathology is the earliest event occurring along the Alzheimer’s *continuum*, which is later followed by tau aggregation and cerebral atrophy (Jack et al., 2016). Both cerebrospinal fluid (CSF) Aβ concentrations and Position Emission Tomography (PET) with specific tracers provide established biomarkers of Aβ pathology. Several previous studies have shown good concordance between these two surrogate markers of Aβ in their diagnostic and prognostic accuracy (Fagan et al., 2006; Landau et al., 2013; Grimmer et al., 2009; Mattsson et al., 2014; Palmqvist et al., 2015). More specifically, a negative relationship between CSF and Aβ PET has been reported both *post-mortem* (Strozyk et al., 2003; Tapiola et al., 2009; La Joie et al., 2019) and *in-vivo* (Toledo et al., 2015; Schindler et al., 2018; Mattsson et al., 2015). They, however, measure two very different pools of Aβ, with CSF Aβ concentrations reflecting the production and clearance rates of soluble Aβ species from the brain, and PET detecting the cumulative load of deposited fibrillary plaques (Roberts et al., 2017; Cohen et al., 2019). It has been suggested that cumulative cerebral Aβ deposition observed in AD might stem from a dysregulation between the production and clearance of Aβ species, and that Aβ plaques may act as a “sink”, hindering the transport of soluble Aβ fragments from the brain to the CSF (Mawuenyega et al., 2010; Blennow et al., 2012). In this respect, the study of factors affecting the balance between soluble and deposited Aβ may help identifying the underlying mechanisms promoting cerebral Aβ aggregation for a given level of CSF Aβ dysmetabolism. With this in mind, we investigated the impact of unmodifiable risk factors known to increase AD prevalence, such as *APOE*-ε4 genotype (Liu et al., 2013), older age (Launer, 2005) and female sex (Ferretti et al., 2018) on the relationship between CSF and Aβ PET markers. We hypothesized that distinct risk factors may exacerbate cerebral Aβ accumulation (assessed by Aβ PET) as a function of incipient Aβ dysmetabolism (assessed by CSF Aβ42/40 concentrations), promoting the formation of fibrillary plaques into specific topological patterns. We tested our hypotheses in regions of vulnerability to AD proteinopathy and further examined the whole-brain using a spatially unbiased voxel-wise approach on a monocentric cohort of middle-aged cognitively unimpaired (CU) participants (ALFA sample). Furthermore, we replicated all analyses in an independent sample of CU participants derived from the Alzheimer’s Disease Neuroimaging Initiative (ADNI).

## METHODS

### Study participants

All participants in the discovery sample were volunteers of the ALFA (ALzheimer and FAmilies) study (Clinicaltrials.gov Identifier: NCT01835717), a longitudinal monocentric research platform aiming at the identification of pathophysiological alterations in preclinical AD. The ALFA cohort entangles 2,743 CU individuals, with a Clinical Dementia Rate score of 0, most of them being first-order descendants of AD patients (Molinuevo et al., 2016). None of the subjects had a neurologic or a psychiatric diagnosis. Within this research framework, the ALFA+ is a nested study that includes advanced imaging protocols, including magnetic resonance imaging (MRI) and PET acquisitions, along with cognitive, lifestyle factors as well as fluid biomarkers. The first 293 consecutive participants of the ALFA+ study with available CSF, Aβ PET, MRI and cognitive data were included in the present work. All the tests and image acquisitions were measured within less than a year time-difference.

The replication sample included all CU ADNI participants (http://adni.loni.usc.edu/), with available Aβ CSF, [^18^F]florbetapir Aβ PET and MRI data acquired within less than one year, resulting in a sample of 259 individuals.

ADNI is a multi-site open access dataset designed to accelerate the discovery of biomarkers to identify and track AD pathology (adni.loni.usc.edu/). The ADNI was launched in 2003 as a public-private partnership, led by Principal Investigator Michael W. Weiner, MD. The primary goal of ADNI has been to test whether serial magnetic resonance imaging (MRI), PET, other biological markers, and clinical and neuropsychological assessment can be combined to measure the progression of mild cognitive impairment (MCI) and early AD. For up-to-date information, see www.adni-info.org. All ALFA participants provided written informed consent and the study was approved by the local ethics committee and conducted according to the principles expressed in the Declaration of Helsinki. Data collection and sharing in ADNI were approved by the Institutional Review Board of each participating institution, and written informed consent was obtained from all participants.

### APOE genotype

For ALFA participants, total DNA was obtained from blood cellular fraction by proteinase K digestion followed by alcohol precipitation. For ADNI, DNA was extracted by Cogenics from a 3-mL aliquot of EDTA blood (adni.loni.usc.edu/data-samples/genetic-data/). Both samples were genotyped for two single nucleotide polymorphisms (SNPs), rs429358 and rs7412, to define the *APOE*-ε2, ε3 and ε4 alleles. For both cohorts, subjects were classified as ε4 carriers (one or two alleles) or non-carriers. Twenty-seven participants in the ALFA cohort being homozygotes for the ε4 allele were excluded from the present study as they were significantly younger than both non-carriers (p<0.001) and *APOE*-ε4 heterozygotes (p<0.001), leading to potential inhomogeneity in the association between CSF and Aβ PET measurements (Rodrigue et al., 2012).

### CSF sampling and analysis

For ALFA participants, CSF samples were obtained by lumbar puncture following standard procedures (Teunissen et al, 2014). CSF was collected into a 15mL sterile polypropylene sterile tube (Sarstedt, Nümbrecht, Germany; cat. no. 62.554.502). CSF was aliquoted in volumes of 0.5mL into sterile polypropylene tubes (0.5mL Screw Cap Micro Tube Conical Bottom; Sarstedt, Nümbrecht, Germany; cat. no. 72.730.005), and immediately frozen at −80°C. Overall, the time between collection and freezing was less than 30 minutes. All the determinations were done in aliquots that had never been previously thawed. Aβ40 and Aβ42 concentrations were determined with the NeuroToolKit (Roche Diagnostics International Ltd) on cobas e 601 (Aβ42) and e 411 (Aβ40) instruments at the Clinical Neurochemistry Laboratory, University of Gothenburg, Sweden. The Roche NeuroToolKit is a panel of exploratory prototype assays designed to robustly evaluate biomarkers associated with key pathologic events characteristic of AD and other neurological disorders. CSF collection and analyses for ADNI participants are described in the ADNI procedure manual (http://adni.loni.usc.edu/methods/). Aβ40 and Aβ42 concentrations in ADNI were measured with 2D-UPLC-tandem mass-spectrometry at the University of Pennsylvania.To increase sensitivity, both in ALFA and ADNI the ratio between Aβ42 and Aβ40 was finally calculated (Lewczuk et al., 2017).

### PET imaging acquisition procedures

Imaging procedures from ALFA have been described previously (Salvadó et al., 2019). In brief, Aβ PET images were acquired 90 min post-injection using [^18^F]flutemetamol with 4 frames of 5 min each. A T1-weighted 3D-TFE sequence was acquired with a 3T Philips Ingenia CX scanner with the following sequence parameters: voxel size = 0.75 mm isotropic, field of view (FOV) = 240 × 240 x 180 mm^3^, flip angle = 8°, repetition time = 9.9 ms, echo time = 4.6 ms, TI = 900 ms.

Details of ADNI imaging procedures can also been found in the website (http://adni.loni.usc.edu/methods/documents/). In brief, [^18^F]florbetapir Aβ PET images were acquired in four frames of five minutes each, 50-70 minutes post-injection. Finally, structural MRI data were acquired on 3T scanning platforms using T1-weighted sagittal 3-dimensional magnetization-prepared rapid-acquisition gradient echo sequences (MP-RAGE).

### Image preprocessing

For both cohorts, individual PET frames were co-registered to produce a mean image, which was subsequently spatially registered onto the respective structural MRI scan. Afterwards, the new segment function in SPM was employed to segment gray matter from MRI scans, which were normalized to the Montreal Neurological Institute (MNI) space, along with the PET images. We calculated the standardized uptake value ratio (SUVR) in MNI space using the whole cerebellum as reference region. Prior to statistical analysis images were smoothed with an 8-mm full width at half-maximum (FWHM) Gaussian kernel.

## Regional Aβ-PET quantification

SUVRs were extracted from a-priori defined regions of interest (ROI). We selected the cortical Centiloid composite ROI (http://www.gaain.org/centiloid-project) as Aβ-sensitive cerebral region (Klunk et al., 2015), which included the following bilateral brain areas: anterior and posterior cingulate cortex, angular gyrus, posterior middle temporal gyrus, middle temporal gyrus, middle frontal gyrus, superior frontal gyrus (pars orbitalis) and the anterior subdivision of the ventral striatum. As tau-vulnerable regions, we selected the Braak stages ROIs (Braak & Braak, 1991) defined according to the Desikan-Killiany atlas (DK atlas) in Schöll et al. (2016). Figure 1 shows both the Centiloid and Braak stages ROIs mapped onto the DK atlas. For visualization purposes, the Centiolid ROIs was parceled onto the DK atlas according to a best-match visual criterion. Supplementary Table 1 shows the full list of the DK atlas labels that were used for both composite ROIs. Supplementary figure 1 shows a surface rendering of the Centilod composite ROI prior to atlas parcellation.

**Fig. 1.**
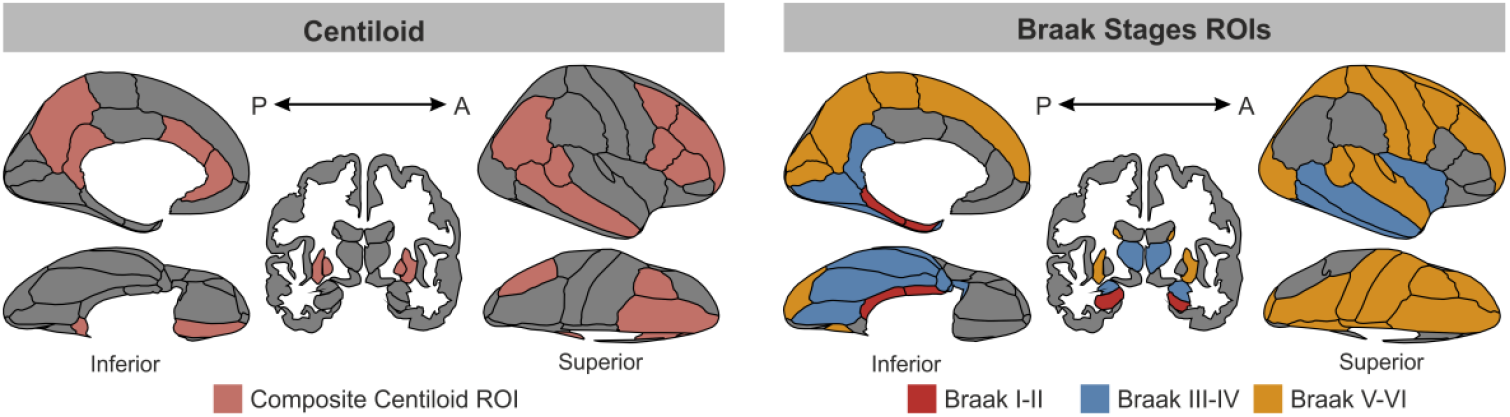
Regions of interest projected onto the Desikan-Killiany atlas. Representation of both cortical and subcortical structures included in the Centiloid and Braak stages ROIs. Polygon rendering is created with the “ggseg” package in R (https://github.com/LCBC-UiO/ggseg/tree/master). A=anterior; P=posterior.

### Neuropsychological assessment

Global cognitive functioning was assessed in both cohorts with the mini mental state examination test (Folstein et al., 1975).

### Statistical analyses

Basic demographic information from both cohorts were compared using *t*-test for continuous variables and Chi-squared test for categorical ones.

We first looked for interactions between CSF Aβ42/40 concentrations and each of the three assessed AD risk factors (*i*.*e*., age, sex and *APOE*-ε4), in promoting cerebral Aβ deposition in regions that are selectively vulnerable to either Aβ (Centiloid composite ROI) or tau pathology (Braak stages ROIs). This first set of analyses was conducted with the SPSS software package (https://www.ibm.com/analytics/spss-statistics-software). Next, we conducted a spatially unbiased whole-brain analysis to detect interaction effects in distributed brain areas. This was achieved by performing a voxel-wise linear regression in SPM12 (Statistical Parametric Mapping, https://www.fil.ion.ucl.ac.uk/spm/). For both the ROI and whole-brain analyses, we set-up three different general linear models where Aβ PET was set as dependent variable, while CSF Aβ42/40, age, sex and *APOE*-ε4 status were modelled as predictors.

Additionally, the interaction term involving CSF Aβ42/40 and any of the three AD risk factors was modelled as independent variable, as follows:

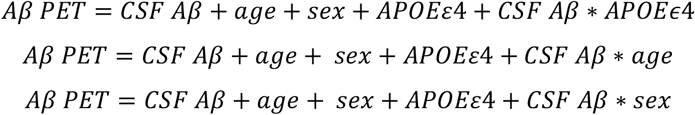

To avoid muticollinearity, continuous CSF Aβ42/40 values were centered to the group mean (Mumford et al., 2015). *APOE*-ε4 was treated as categorical binary variable (*i*.*e*., 0=non-carriers, 1=ε4-carriers). In SPM, we set parametric t-contrasts on the interaction terms, based on the hypothesis that each risk factor would exacerbate amyloid fibrillary deposition as function of CSF Aβ42/40 concentrations.

Finally, to assess the combined effects of AD risk factors, we performed additional analyses testing three-way interactions involving CSF Aβ42/40 and each pair of the tested risk factors. To this aim, we set up three different statistical models where the effects of CSF Aβ42/40 on Aβ PET were studied in combination with either *APOE*-ε4 and age, *APOE*-ε4 and sex, or age and sex.

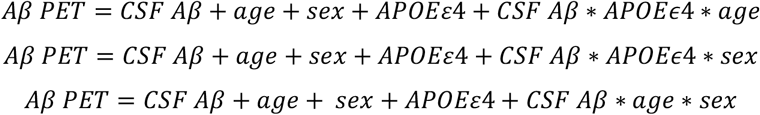

For the ROI analyses, results were considered significant if surviving a threshold of p<0.05 corrected for multiple testing using a False-Discovery Rate (FDR) approach. For the whole brain voxel-wise analysis, we set a threshold of p<0.001 and applied a cluster extent correction of 100 contiguous voxels (k > 100). All the above-mentioned statistical models were applied to the ADNI replication sample. Voxel-wise analyses in ADNI were masked with an inclusive mask derived from the analyses conducted in ALFA, which was generated at a liberal threshold of p<0.005 with a cluster extent of 100 voxels.

## RESULTS

### Sample characteristics

Demographic characteristics of both cohorts can be found in Table 1. Compared to ALFA, ADNI participants were significantly older, more educated, and harbored a lower proportion of *APOE*-ε4 carriers. However, the two samples were homogeneous with respect to sex and global cognitive performance. As expected, in both cohorts CSF Aβ42/40 concentrations were negatively related to Aβ PET uptake in widespread cortical areas, while age and *APOE*-ε4 were positively associated to cortical Aβ deposition (Supplementary Figure 2).

**Table 1.**
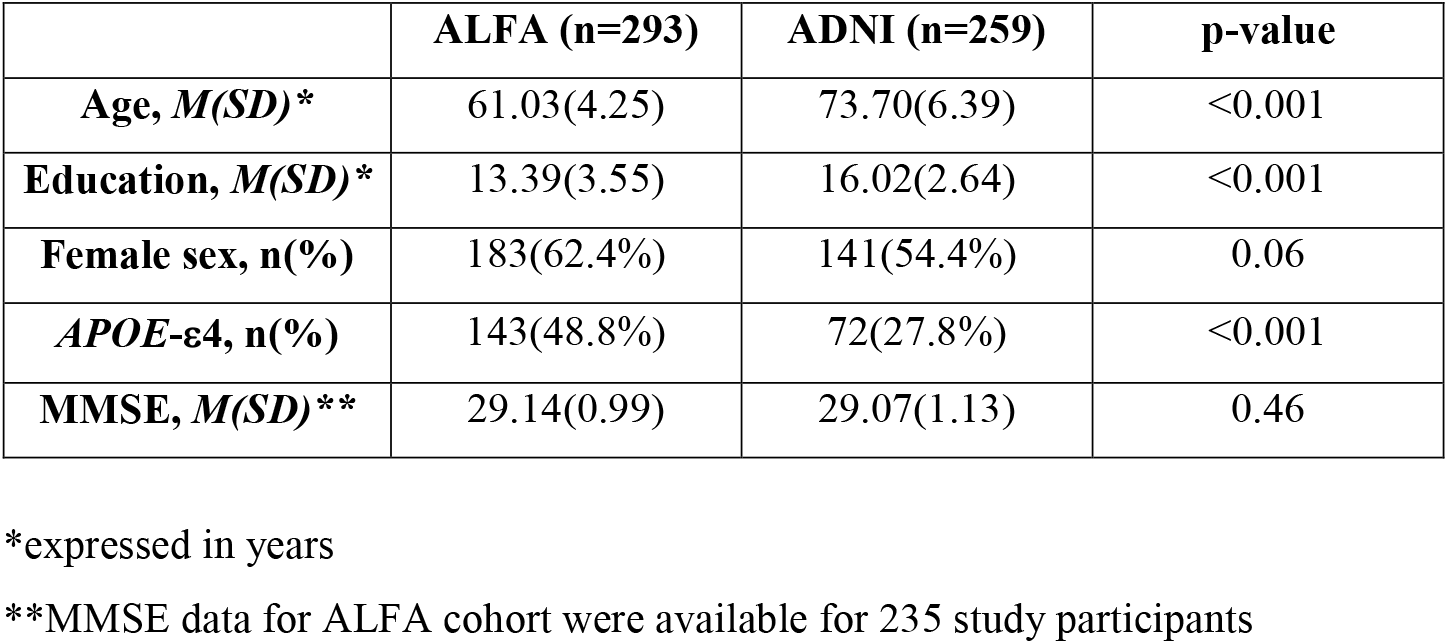
Sample characteristics.

### ROI analyses

We assessed whether AD risk factors such as age, *APOE*-ε4 and female sex modulated the association between soluble and deposited Aβ in cerebral regions known for their vulnerability to either Aβ or tau pathology. Table 2 shows the results of each statistical model run for the different risk factors in each of the tested ROIs; for each model, the F-statistic and p-value of the interaction term are presented. Within the Centiloid ROI, each of the 2-Way and 3-Way interactions were significant in the ALFA cohort, while in ADNI the interaction between CSF Aβ42/40 and sex did not survive statistical correction. In Braak I/II ROIs, only the 2-Way and 3-Way interactions involving *APOE*-ε4 were significant in ALFA but not the remaining models, while in ADNI no significant interactions were found. In Braak III/IV ROIs, we observed similar results as for the Centiloid ROI, that is, all interaction models being significant in ALFA with two interactions not reaching statistical significance in ADNI, those between CSF Aβ42/40 and sex as well as CSF Aβ42/40 and *APOE*-ε4. Finally, in Braak V/VI, ALFA participants displayed all significant interactions except a statistical trend for CSFAβ42/40 x *APOE*-ε4, while for ADNI participants only the two way interaction involving sex did not reach statistical significance. Figure 2 shows group scatterplots highlighting the modulatory role of each risk factor on the association between soluble and deposited Aβ in those ROIs, for both cohorts.

**Table 2.** **Interactions between CSF Aβ_42/40_ and each AD risk factors on Aβ-PET uptake in vulnerable ROIs**

|  | Centiloid ROI |  |  |  | Braak I/II |  |  |  | Braak III/IV |  |  |  | Braak V/VI |  |  |  |
| --- | --- | --- | --- | --- | --- | --- | --- | --- | --- | --- | --- | --- | --- | --- | --- | --- |
|  | ALFA |  | ADNI |  | ALFA |  | ADNI |  | ALFA |  | ADNI |  | ALFA |  | ADNI |  |
|  | F<sub>1,287</sub> | pFDR | F<sub>1,251</sub> | pFDR | F<sub>1,287</sub> | pFDR | F<sub>1,251</sub> | pFDR | F<sub>1,287</sub> | pFDR | F<sub>1,251</sub> | pFDR | F<sub>1,287</sub> | pFDR | F<sub>1,251</sub> | pFDR |
| CSFA $\beta$ * *APOE* - $\epsilon 4$ | 6.89 | **0.013** | 6.21 | **0.024** | 12.73 | **0.001** | 0.178 | 0.702 | 10.47 | **0.002** | 2.80 | 0.142 | 3.58 | 0.071 | 4.83 | **0.049** |
| CSFAβ*age | 36.40 | **<0.001** | 8.62 | **0.011** | 1.28 | 0.269 | 4.44 | 0.062 | 27.48 | **<0.001** | 10.74 | **0.008** | 29.01 | **<0.001** | 7.79 | **0.014** |
| CSFAβ*sex | 5.51 | **0.027** | 0.72 | 0.453 | 1.44 | 0.252 | 0.65 | 0.459 | 6.64 | **0.037** | 0.58 | 0.810 | 5.07 | **0.032** | 1.12 | 0.348 |
| CSFA $\beta$ *age* *APOE* - $\epsilon 4$ | 22.47 | **0.002** | 6.20 | **0.002** | 8.48 | **<0.001** | 1.71 | 0.219 | 20.20 | **<0.001** | 5.20 | **0.010** | 16.61 | **<0.001** | 5.38 | **0.012** |
| CSFA $\beta$ *sex* *APOE* - $\epsilon 4$ | 5.36 | **0.002** | 3.75 | **0.010** | 5.14 | **0.003** | 1.49 | 0.244 | 6.69 | **<0.001** | 3.09 | **0.020** | 3.98 | **0.012** | 3.21 | **0.017** |
| CSFAβ*age*sex | 21.72 | **0.001** | 5.98 | **0.012** | 1.31 | 0.272 | 2.24 | 0.162 | 16.76 | **0.001** | 6.35 | **0.012** | 17.69 | **<0.001** | 5.90 | **0.009** |

CSF: Cerebrospinal fluid; pFDR: p-value adjusted for multiple testing with False Discovery Rate

**Fig. 2.**
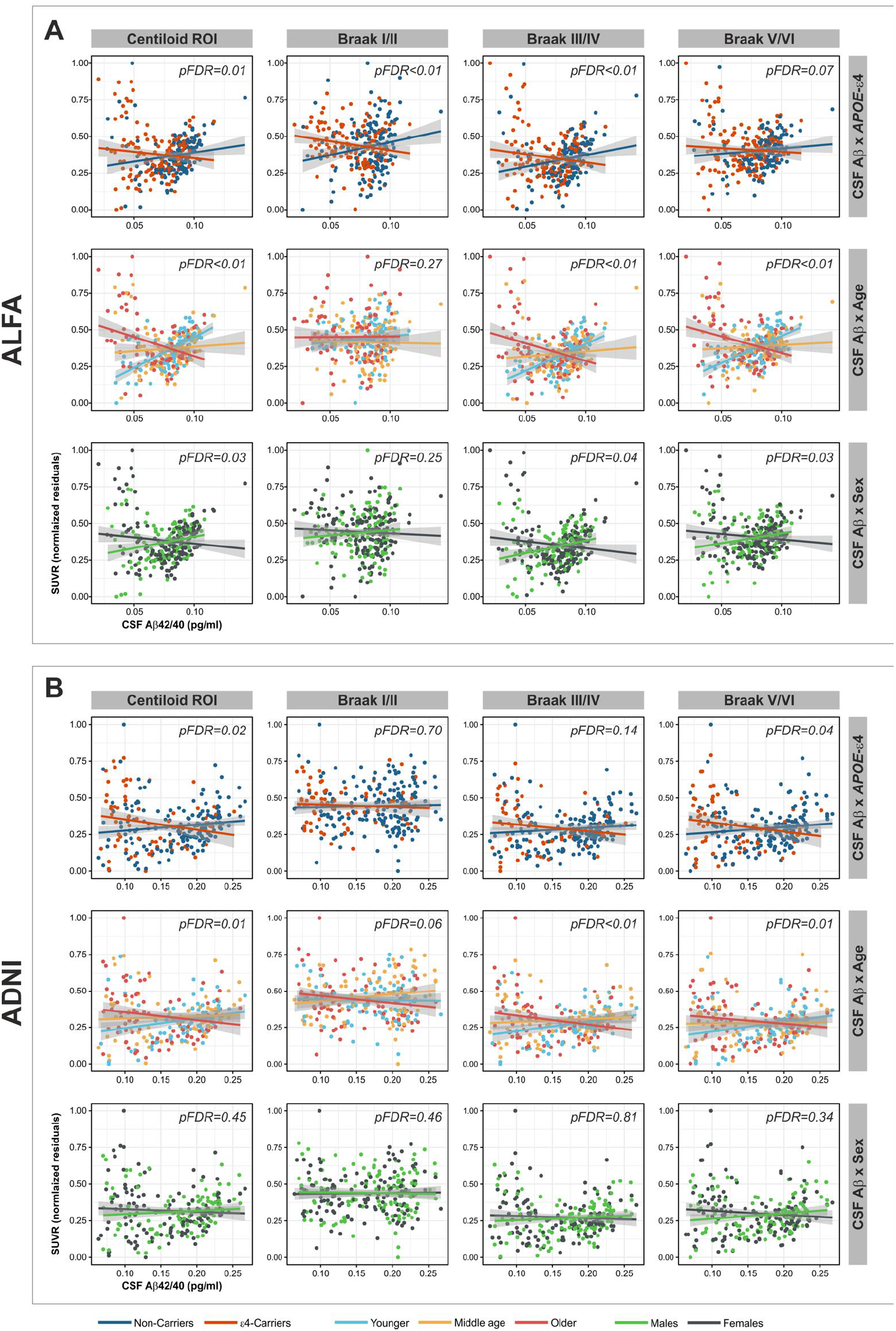
*APOE*-ε4, age and sex modified the association between CSF Aβ42/40 concentrations and Aβ PET uptake quantified in regions of interest·. **A)** Interactions assessed in the ALFA sample and **B)** in ADNI. SUVRs were residualized against the covariates of interest in each model (refer to the Statistical Analysis section). For visualization purposes, age continuous variable was broken down in three subgroups of younger, middle-aged, and older individuals, according to tercile ranking.

### Whole brain analysis: two-way interactions

We then examined the impact of each risk factor on the association between soluble and deposited Aβ on the whole brain level by conducting voxel-wise regressions. In the ALFA cohort, we found that, for any given value of CSF Aβ42/40, *APOE*-ε4 carriers displayed a higher Aβ PET retention in the bilateral anterior hippocampus extending to the entorhinal cortex, also including the inferior temporal and angular gyrus bilaterally (Fig. 3a-3c). These results were replicated in the ADNI cohort, whereby the interaction between CSF Aβ42/40 and *APOE*-ε4 was significant in a topographical pattern consistent with that of ALFA, and including the right middle and inferior temporal cortex, as well as the anterior cingulate cortex (ACC) and right angular gyrus (Fig. 3d-3f).

**Fig. 3.**
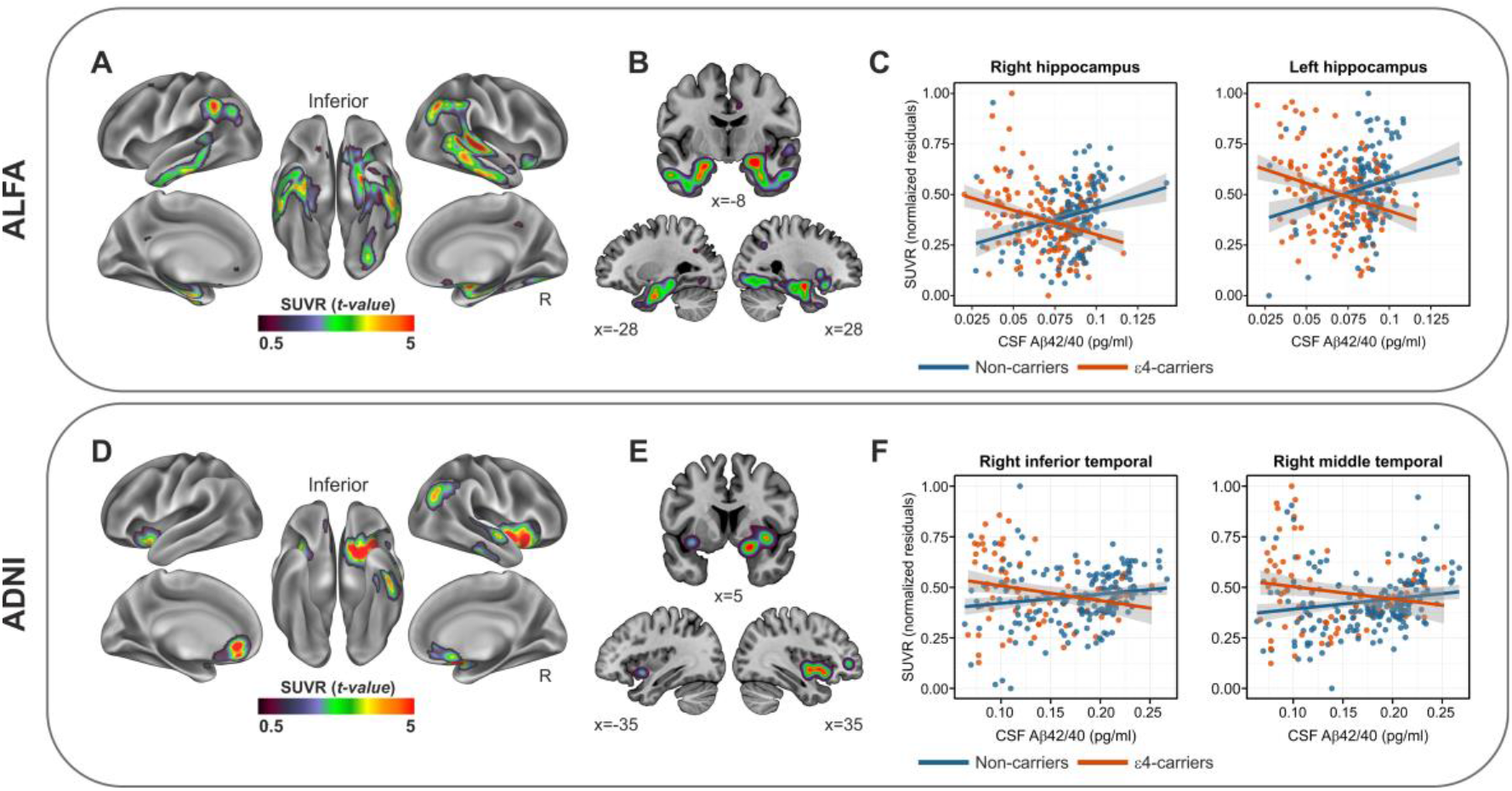
*APOE*-ε4 significantly modified the spatial topography of Aβ PET as function of CSF Aβ42/40·. **A-B)** Surface and volume rendering in ALFA participants of the Aβ PET statistical t-map resulting from the interaction model. Data indicate that compared to non-carriers, *APOE*-ε4 carriers displayed higher SUVRs, for any given level of CSF Aβ42/40, in medial temporal regions including entorhinal cortex and hippocampus. **C)** Group scatterplots in ALFA participants showing the significant interaction between *APOE*-ε4 and CSF Aβ42/40 in driving Aβ PET SUVRs in the right and left hippocampus. **D-E)** Surface and volume rendering in ADNI participants, of Aβ PET statistical t-map indicating that compared to non-carriers, *APOE*-ε4 carriers displayed higher SUVRs, for any given level of CSF Aβ42/40, in right inferior and middle temporal as well as right insula. **F)** Group scatterplots in ADNI participants showing the significant interaction between *APOE*-ε4 and CSF Aβ42/40 in driving Aβ PET SUVRs in the right inferior and middle temporal gyrus.

Next, we found that age modulated the association between CSF Aβ42/40 concentration and cortical Aβ deposition, with older individuals displaying greater SUVRs in bilateral superior frontal cortex, middle and inferior temporal areas as well as anterior and posterior cingulate cortex (PCC) (Fig. 4a-4b) in the ALFA cohort. Such a pattern of results was replicated in ADNI, with similar effects shown in older compared to younger individuals, even though the topological pattern was less widespread (Fig 4c-4d).

**Fig. 4.**
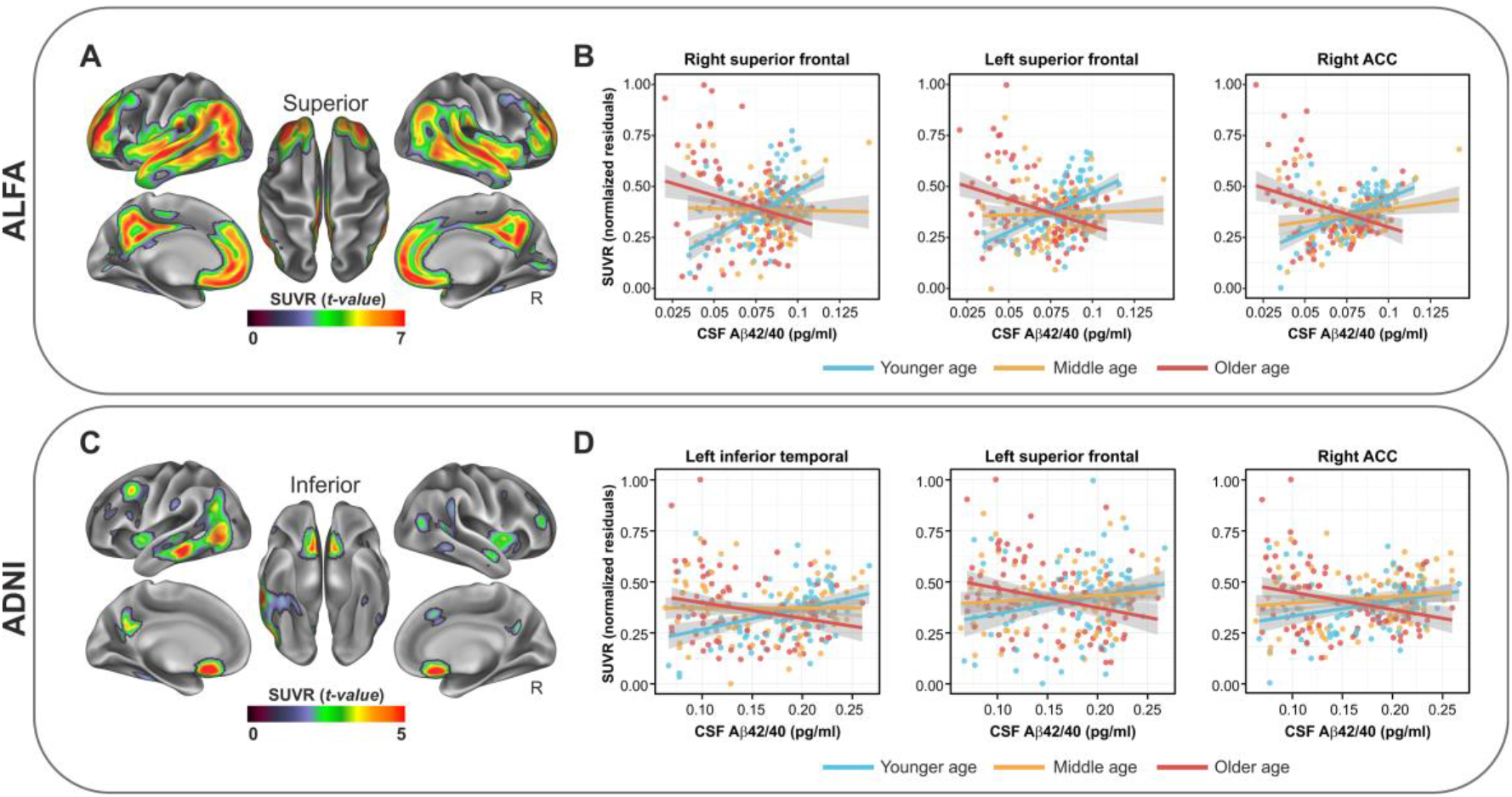
Age significantly modified the association between of CSF Aβ42/40 and Aβ PET. **A-B)** In ALFA participants, older individuals displayed, for any given level of CSF Aβ42/40 concentration, a higher Aβ PET retention in distributed cerebral areas including inferior and superior temporal cortex as well as medial prefrontal and inferior parietal areas. **C-D)** Such an interaction was replicated in the ADNI cohort, although in a less distributed topological pattern. For visualization purposes, age continuous variable was broken down in three subgroups of younger, middle-aged, and older individuals, according to tercile ranking.

Finally, a similar modulatory role was observed for sex, indicating higher SUVRs in posterior areas including the cuneus, the middle cingulate cortex and middle temporal gyrus, in females compared to males (Fig. 5a-5b) in the ALFA cohort. As for the previous interactions, this interaction was replicated in ADNI, even though the spatial topography was less distributed, including the cuneus bilaterally, as shown in Fig 5c-5d.

**Fig. 5.**
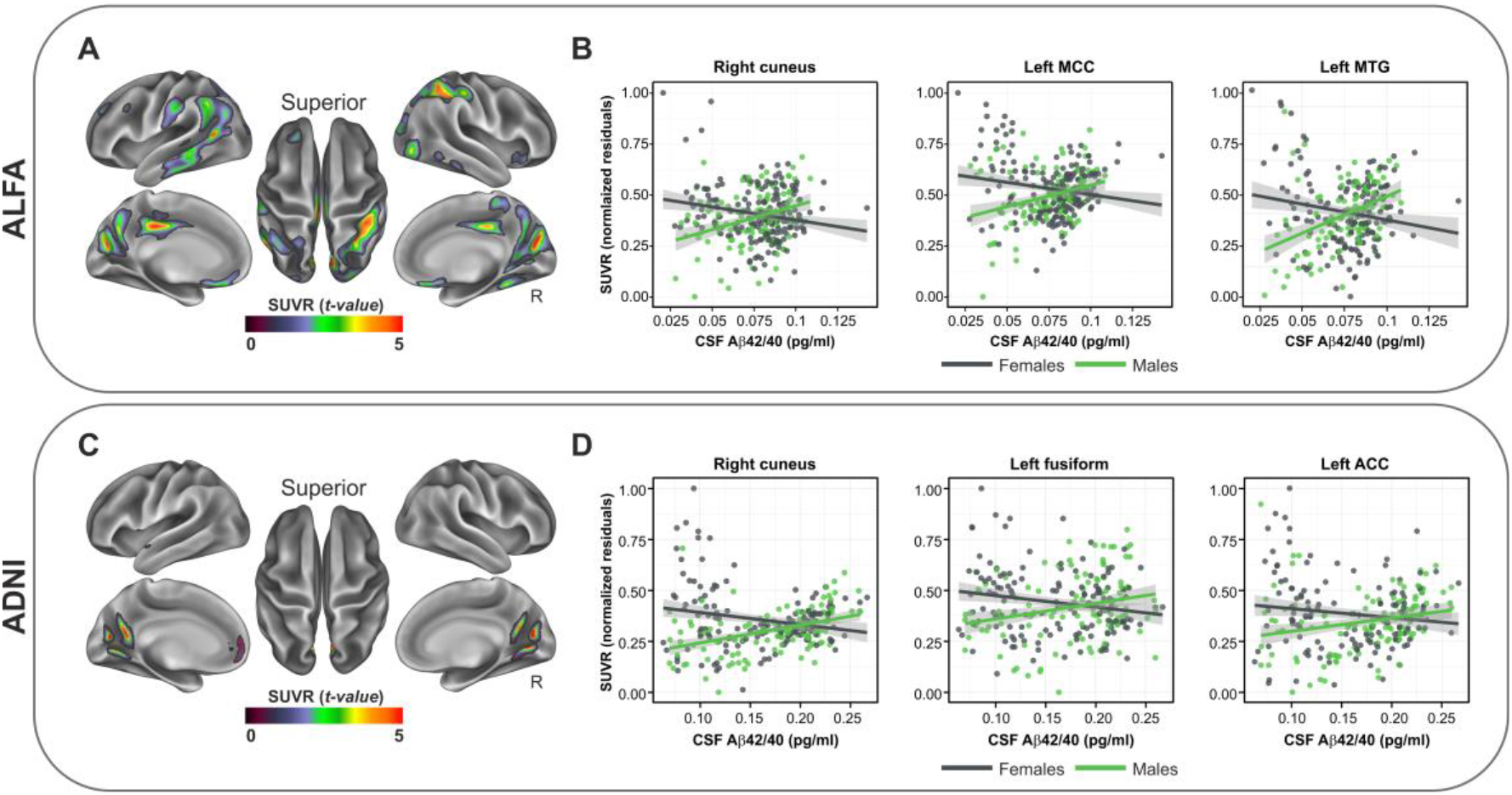
Sex significantly modified the association between CSF Aβ42/40 and Aβ PET. **A-B)** In ALFA participants, sex significantly modulated the association between CSF Aβ42/40 and Aβ PET uptake indicating higher SUVRs in females participants compared to males, in posterior medial regions including the precuneus and the cuneus. **C-D)** An overlapping cortical topology was found in ADNI participants indicating the same interaction effects as in ALFA. rITG=right inferior temporal gyrus; lITG=left inferior temporal gyrus; rPUT

### Whole brain voxel-wise analysis on the synergistic effects of APOE-ε4, age and sex

In the ALFA cohort, we observed a significant three-way interaction involving CSF Aβ42/40, *APOE*-ε4, and age in lateral temporal regions, temporo-parietal junction, and PCC, indicating that the detrimental effects of *APOE*-ε4 in driving a higher Aβ PET uptake as function of CSF Aβ42/40 concentrations, were stronger in older compared to younger individuals (Fig. 6a-6b). Similar patterns were significant in ADNI (Fig. 6c-6d). Next, we found a three-way interaction involving CSF Aβ42/40, age and sex, indicating that the modulatory role of age in prompting a higher Aβ PET uptake as function of CSF Aβ42/40 concentrations, were stronger in female compared to male individuals. This interaction mapped onto the orbitofrontal cortex, inferior parietal as well as anterior and posterior cingulate (Fig. 6e-6f). These findings, although less widespread, were replicated in ADNI participants as well (Fig. 6g-6h).

**Fig. 6.**
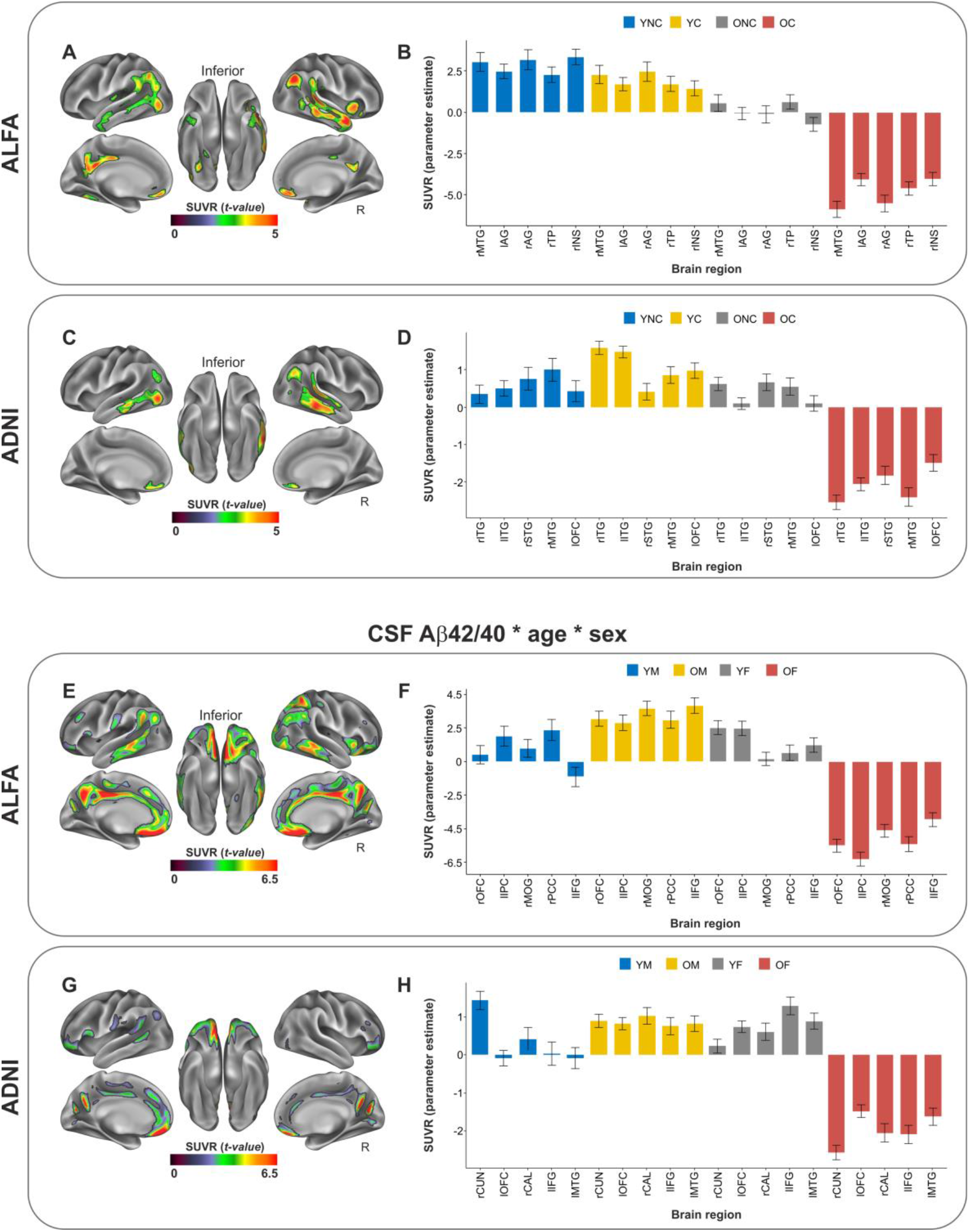
Three-way interactions in both cohorts. **A-B)** Surface rendering and bar plots indicating three-way interactions involving CSF Aβ42/40, *APOE*-ε4 and age on Aβ PET uptake in the ALFA cohort. YNC=young non-carriers; YC=young ε4-carriers; ONC=older non-carriers; rMTG=right middle temporal gyrus; lAG=left angular gyrus; rAG=right angular gyrus; rTP=right temporal pole. **C-D)** Same as in A-B, in the ADNI cohort. rINS=right insula; rITG=right inferior temporal gyrus; lIPG=left inferior parietal gyrus; rSTG=right superior temporal gyrus; lOFC=orbitofrontal cortex. In B) and D), bars in the plot encode the interaction between one categorical (*APOE*-ε4) and two continuous (CSF Aβ42/40, age) variables. **E-F)** Surface rendering and bar plots indicating three-way interactions involving CSF Aβ42/40, age and sex on Aβ PET uptake in the ALFA cohort. YM=young males; OM=older males; YF=young females; OF=older females; rOFC=right orbitofrontal cortex; lIPC=left inferior parietal cortex; rMOG=right medial orbital gyrus; rPCC=right posterior cingulate cortex; lIFG=left inferior frontal gyrus; **G-H)** Same as in E-F, in the ADNI cohort. rCUN=right cuneus; lOFC=left orbitofrontal cortex; rCAL=right calcarine; lIFG=left inferior frontal gyrus; lMTG=left middle temporal gyrus. In F) and H), bars in the plot encode the interaction between one categorical (*s*ex) and two continuous (CSF Aβ42/40, age) variables.

Finally, we found a significant three-way interaction involving CSF Aβ42/40, *APOE*-ε4, and sex, indicating that the exacerbating effects of *APOE*-ε4 were more prominent in women compared to men, in inferior temporal and orbitofrontal regions (Fig. 7a-7b). This interaction was however not replicated in ADNI.

**Fig. 7.**
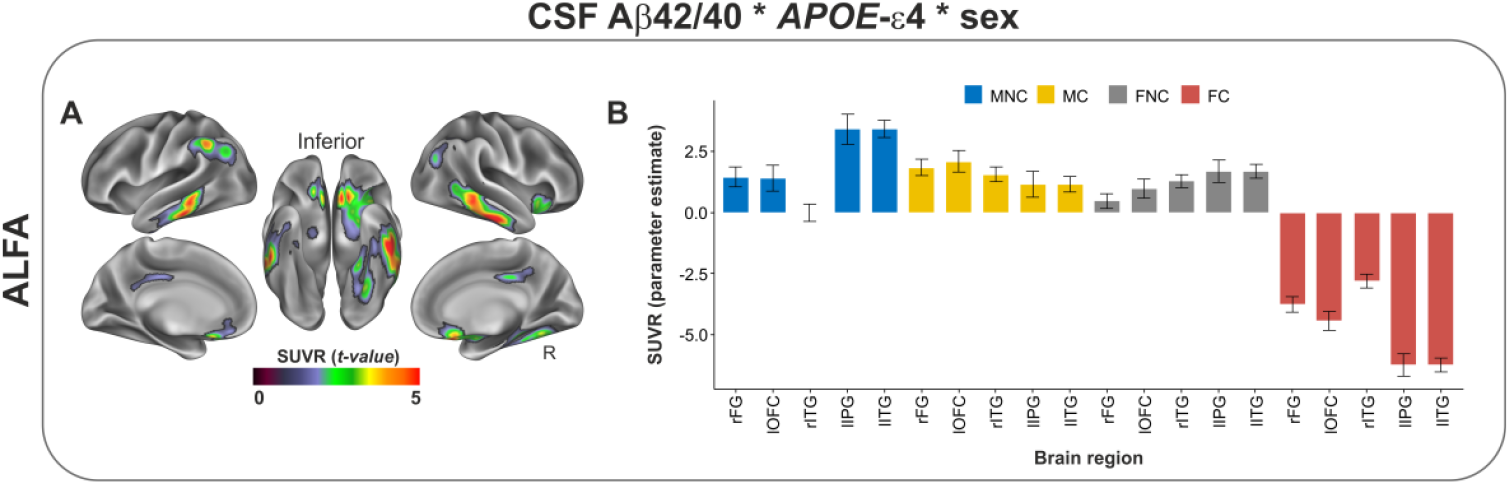
Three-way interactions in the ALFA cohort. **A-B)** Surface rendering and bar plots indicating three-way interactions involving CSF Aβ42/40, age and sex on Aβ PET uptake in the ALFA cohort. MNC=males non-carrier; MC=males ε4-carriers; FNC=females non-carrier; FC=females ε4-carriers; FG=right fusiform gyrus; lOFC=orbitofrontal cortex; rITG=right inferior temporal gyrus; lIPG=left inferior parietal gyrus; lITG=left inferior temporal gyrus. Bars in the plot encode the interaction between two categorical (*APOE*-ε4, sex) and one continuous (CSF Aβ42/40) variables.

## DISCUSSION

The present work aimed to determine whether in CU individuals, unmodifiable AD risk factors modulate the association between soluble and deposited Aβ species quantified with CSF concentrations and PET imaging, respectively. Further, our goal was to determine which brain regions are susceptible for such differential associations. We found that *APOE*-ε4, older age and female sex, all interacted with CSF Aβ42/40 concentrations, resulting in a higher fibrillary plaque deposition for any given level of CSF Aβ42/40, with each risk factor mapping onto a specific topology. Importantly, we replicated these findings in an independent cohort that differed in the PET tracers used for Aβ imaging as well as in the average age and level of progression in the preclinical AD *continuum*, thus reinforcing the robustness and generalizability of our results. Our strategy of assessing the impact of risk factors on the association between two distinct surrogate markers of cleared and aggregated Aβ in CU individuals provide novel insights into the biological pathways underlying Aβ aggregation in the brain.

First, we observed a significant interaction between CSF Aβ42/40 and *APOE*-ε4 in the Centiloid ROI as well as in Braak stages I/II and III/IV regions, in the ALFA cohort. In ADNI, this interaction was significant in the Centiloid ROI, as well as the Braak stages V/VI ROI. Whole-brain analyses conducted in the ALFA cohort confirmed that, compared to non-carriers, *APOE*-ε4 carriers displayed, for any given value of CSF Aβ42/40, a higher Aβ PET retention in a symmetric pattern covering medial temporal lobe (MTL) areas including the anterior hippocampus, parahippocampus, entorhinal cortex, inferior temporal as well as the bilateral inferior parietal regions. Similarly, in ADNI this interaction covered the right middle and inferior temporal cortex, as well as the anterior cingulate cortex and right angular gyrus. These areas do not typically display Aβ accumulation in the early stages of the disease, which rather involve neocortical areas and particularly prefrontal cortex, posterior cingulate, precuneus, and inferior parietal, as shown by *in-vivo* staging (Collij et al., 2020; Mattsson et al., 2019; Grothe et al., 2017) and autopsy studies (Braak & Braak, 1991; Thal et al., 2002). Rather, the regions we found, particularly in the ALFA sample, display selective vulnerability to early tau deposition, as previously documented in patients along the Alzheimer’s *continuum* (Ossenkoppele et al., 2016; Schwarz et al., 2016; Cho et al., 2016) as well as in cognitively unimpaired individuals (Johnson et al., 2016; Schöll et al., 2016). Earlier studies provide evidence for a synergistic interaction between Aβ and tau in determining functional and structural abnormalities in cognitively intact individuals (Busche & Hyman, 2020; Pascoal et al., 2017; Desikan et al., 2012; Fortea et al., 2014). According to a disease model of Aβ-induced tau hyperphosphorylation (Maia et al., 2013; Schelle et al., 2017), fibrillary Aβ initiates a pathophysiological cascade leading to tau misfolding that eventually propagates throughout the neocortex. Furthermore, one study reported that the interaction between Aβ and tau in driving a greater risk of developing AD, mapped onto inferior temporal and parietal regions, which overlap with the regions we found (Pascoal et al., 2017). Hence, our results suggest that *APOE*-ε4, by affecting the association between soluble and deposited Aβ specifically in MTL areas, paves the way for the spread of tau in extra medial-temporal regions thus promoting later co-localization of Aβ and tau. Indeed, previous PET imaging studies have documented a higher tau deposition in *APOE*-ε4 carrier AD patients compared to non-carriers (Ossenkoppele et al., 2016; Therriault et al., 2020; Tiraboschi et al., 2004). The importance of this co-localization is highlighted by evidence that the presence of tau pathology beyond the mesial temporal lobe is facilitated by the presence of Aβ in these regions (Pontecorvo et al., 2017; Jones et al., 2017; He et al., 2018; Vogel et al., 2020; Shimada et al., 2017). In turn, tau deposition in MTL regions drives subsequent neurodegeneration, brain atrophy and cognitive decline (La Joie et al., 2020; Bejanin et al, 2017). These regional effects were not detected for the interactions between CSF Aβ42/40 and age or sex, but post-hoc three-way interactions indicated that the effect of *APOE*-ε4 was significantly stronger for females and older participants. Our data support earlier evidence for a combined influence of age, sex and *APOE*-ε4 on the emergence of AD pathology (Mofrad et al., 2020; Li et al., 2017; Lautner et al,. 2017; Glodzik-Sobanska et al., 2009), and AD prevalence (Riedel et al., 2016; Raber et al., 2004; Farrer et al, 1997; Jarvik et al., 1995).

Furthermore, our interaction effects may help to explain the faster disease progression (Mishra et al., 2018; Paranjpe et al., 2019) as well as the stronger relationship between Aβ and cognitive decline (Mormino et al., 2014; Kantarci et al., 2012; Lim et al., 2015) in *APOE*-ε4 carriers compared to non-carriers. It is worth noting that, when assessing the main effects of *APOE*-ε4 on Aβ PET we found, as expected, a widespread higher retention in ε4-carriers compared to non-carriers across regions consistent with those reported previously, namely prefrontal and midline cortical areas (Supplementary Figure 2) (Reiman et al., 2009; Toledo et al., 2019). However, as mentioned above, our interaction data between CSF Aβ42/40 and *APOE*-ε4 revealed a cortical topology in different areas and precisely in MTL, tau-vulnerable, regions. Thus, our results imply that the ratio between cerebral deposited Aβ and its soluble counterpart may represent a novel biomarker putatively reflecting the imbalance between cleared and deposited Aβ in the brain, and may thus be more informative on the mechanisms of incipient Aβ pathology. Further longitudinal studies are required to track the progression of regional Aβ PET uptake as function of CSF Aβ concentrations stratified by genetic risk, age and sex.

It is important to note that the ALFA and ADNI cohorts had significantly different age (mean age ALFA: 61.03; mean age ADNI: 73.71) and levels of cerebral Aβ load (mean Centiloid in ALFA = 2.66; mean Centiloid in ADNI = 25.71), which can be regarded as a proxy for disease progression (Palmqvist et al., 2019). Therefore, the comparison of the effect sizes across cohorts in progressive Braak stages ROIs may be indicative of the timing within the disease continuum when the effect of each of these AD risk factors may be more relevant in promoting the accumulation of Aβ in tau-vulnerable regions. In general, the sizes of effects of these AD risk factors in ADNI were progressively larger in Braak V/VI than in Braak III/IV, and nonsignificant in Braak I/II. In contrast, in ALFA, whose participants are supposed to be in earlier stages of the Alzheimer’s *continuum*, the interaction effect between CSF Aβ and *APOE*-ε4 was mostly prominent in Braak I/II, followed by Braak III/IV. This pattern suggests that the effect of *APOE*-ε4 in promoting Aβ aggregation in entorhinal regions, which is thought to be a key event in the development of AD, may happen very early in the AD *continuum*.

Next, we reported a modulatory effect for older age in driving a higher Aβ PET uptake as a function of CSF Aβ42/40. This interaction was significant in the Centiloid ROI, Braak stages III/IV and stages V/VI, in both cohorts. Whole brain analyses yielded significant effects into the posterior cingulate, precuneus, medial prefrontal cortex including the ACC, as well as superior frontal and middle temporal areas in the ALFA cohort. Consistent effects were found in the ADNI sample even though within a more restricted topology involving ACC, PCC and middle temporal areas. Such a reduced effect in ADNI compared to that in ALFA may be due to the different age range of the two samples (ALFA= 50-73; ADNI=56-94 years). In fact, earlier studies indicate that, the effects of aging on cortical Aβ deposition drop significantly in cognitively intact individuals older than 60 years of age (*e*.*g*., Rodrigue et al., 2012). Unlike the interaction findings involving *APOE*-ε4, the brain regions susceptible for the interaction between CSF Aβ42/40 and age display selective vulnerability to Aβ accumulation early in the disease *continuum* (Collij et al., 2020; Mattsson et al., 2019; Braak & Braak, 1991; Thal et al., 2002). These regions overlap with the default-mode network, which is known for harboring Aβ in the initial stages of the disease, as well as in normal aging (Buckner et al., 2005, 2009; Palmqvist et al,. 2017). Interestingly, aging has been associated to a progressive disruption of the DMN, which in turn affects memory efficiency (Damoiseaux et al., 2008; Miller et al, 2008). Hence, our interaction data suggest that, as Aβ clearance rate begins to become deficient, older individuals harbor more fibrillary plaques within the DMN, which may exacerbate the effects of Aβ on cognitive performance. This hypothesis however goes beyond the scope of the present study.

Finally, we reported that sex modulated the association between soluble and deposited Aβ in the Centiloid ROI, as well as Braak stages ROIs III/IV and V/VI in the ALFA cohort, while no significant two-way interactions involving sex were retrieved in the ADNI sample using an ROI approach. Whole brain analyses yielded a significant interaction in posterior medial regions such as the PCC and cuneus, as well as middle temporal areas in ALFA, while a less distributed effect was found in ADNI, involving the cuneus bilaterally. Women represent two-thirds of the cases of late-onset AD (Beam et al., 2018). Such a higher prevalence may be ascribed to several factors, including sex-related differences in neuroinflammation burden (Hall et al., 2013), higher proportion of major depressive disorders in women (Albert, 2015) and most importantly, estrogen depletion occurring in the perimenopause (Brinton et al., 2015). Reductions in estrogen levels in women during the fifth decade and beyond, may be responsible for deficits in the brain metabolism and vascular pathogenesis. Compared to perimenopausal, post-menopausal women show more prominent brain hypometabolism, increased Aβ deposition and reduced gray matter volume, with these effects mapping onto posteromedial cortices (Mosconi et al., 2017), similarly to the brain areas that we have found in the interaction analysis. In addition, sex has been shown to modify the *APOE*-related increased risk of developing AD, with a higher proportion of MCI to AD converters in women than in men ε4-carriers (Altman et al., 2014). Our three-way interaction showing a greater deleterious effect of *APOE*-ε4 on Aβ deposition in women than in men suggest that Aβ-dependent mechanisms may underlie those previous observations. More in general, our three way interaction results indicate that cognitively intact older females harboring *APOE*-ε4 may enter earlier in preclinical AD stage, thus calling for their inclusion in primary prevention strategies.

In conclusion, we show that *APOE*-ε4, age and sex modify the relationship between soluble and deposited Aβ, with each risk factor promoting a higher cerebral Aβ deposition as function of CSF Aβ42/40 concentrations in specific brain regions. The interaction between CSF Aβ and *APOE*-ε4 mapped onto regions that do not typically accumulate Aβ in the early preclinical AD stages, but rather show selective vulnerability to tau aggregation and atrophy early in the disease. On the other hand, both age and sex interactions showed their effects on areas of initial Aβ deposition. These data provide novel insights into the factors mediating the balance between soluble and deposited Aβ, and clarify the mechanisms underlying the higher AD prevalence associated to those risk factors.

## Data Availability

Data used in preparation of this article were obtained from the Alzheimer's Disease Neuroimaging Initiative (ADNI) database (adni.loni.usc.edu). As such, the investigators within the ADNI contributed to the design and implementation of ADNI and/or provided data but did not participate in analysis or writing of this report. A complete listing of ADNI investigators can be found at: http://adni.loni.usc.edu/wp-content/uploads/how_to_apply/ADNI_Acknowledgement_List.pdf

http://adni.loni.usc.edu

## Acknowledgements

This publication is part of the ALFA study (ALzheimer and FAmilies). The authors would like to express their most sincere gratitude to the ALFA project participants, without whom this research would have not been possible. Authors would like to thank Roche Diagnostics International Ltd. for kindly providing the kits for the CSF analysis of ALFA+ participants and GE Healthcare for kindly providing [^18^F]flutemetamol doses of ALFA+ participants. COBAS and COBAS E are registered trademarks of Roche. The project leading to these results has received funding from “la Caixa” Foundation (ID 100010434), under agreement LCF/PR/GN17/50300004 and the Alzheimer’s Association and an international anonymous charity foundation through the TriBEKa Imaging Platform project (TriBEKa-17-519007). Additional support has been received from the Universities and Research Secretariat, Ministry of Business and Knowledge of the Catalan Government under the grant no. 2017-SGR-892. JDG is supported by the Spanish Ministry of Science and Innovation (RYC-2013-13054). MSC receives funding from Instituto de Salud Carlos III (PI19/00155) and from the Spanish Ministry of Science, Innovation and Universities (Juan de la Cierva programme grant IJC2018-037478-I). KB holds the Torsten Söderberg Professorship in Medicine at the Royal Swedish Academy of Sciences, and is supported by the Swedish Research Council (#2017-00915); the Swedish Alzheimer Foundation (#AF-742881), Hjärnfonden, Sweden (#FO2017-0243); and a grant (#ALFGBG-715986) from the Swedish state under the agreement between the Swedish government and the County Councils, the ALF-agreement. HZ is a Wallenberg Scholar supported by grants from the Swedish Research Council (#2018-02532), the European Research Council (#681712), Swedish State Support for Clinical Research (#ALFGBG-720931), the Alzheimer Drug Discovery Foundation (ADDF), USA (#201809-2016862), the European Union’s Horizon 2020 research and innovation programme under the Marie Skłodowska-Curie grant agreement No 860197 (MIRIADE), and the UK Dementia Research Institute at UCL.

Collaborators of the ALFA study are: Müge Akinci, Annabella Beteta, Alba Cañas, Irene Cumplido, Carme Deulofeu, Ruth Dominguez, Maria Emilio, Karine Fauria, Sherezade Fuentes, Oriol Grau-Rivera, José M. González de Echevarri, Laura Hernandez, Gema Huesa, Jordi Huguet, Iva Knezevic, Eider M. Arenaza-Urquijo, Eva M Palacios, Paula Marne, Tania Menchón, Carolina Minguillon, Albina Polo, Sandra Pradas, Blanca Rodríguez, Aleix Sala-Vila, Gonzalo Sánchez-Benavides, Anna Soteras, Laura Stankeviciute, Marc Vilanova and Natalia Vilor-Tejedor.

