## Supplementary Materials for "Age, sex and *APOE*-ε4 modify the balance between soluble and fibrillar β-amyloid in cognitively intact individuals: topographical patterns and replication across two independent cohorts"

### *Supplementary statistical analyses*

In both the ALFA and ADNI cohorts, main effects of CSF A$\beta_{42/40}$, age, sex, and *APOE*-ε4 on Aβ-PET uptake were assessed by setting up a voxel-wise linear regression in SPM12 (https://www.fil.ion.ucl.ac.uk/spm/ ), with the normalized and smoothed SUVR individual parametric maps entered as dependent variables, while modeling the above mentioned factors as independent explanatory variables. Results were considered significant if surviving a voxel-level threshold of $p<0.001$ applying a cluster extent correction of 100 contiguous voxels ($k > 100$).

### *Centiloid composite region of interest (ROI)*

Fig. S1 shows a surface rendering of the Centiloid composite ROI.

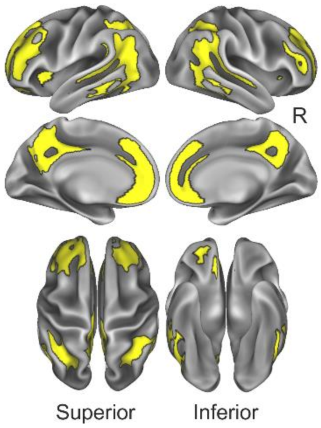


**Supplementary figure 1** – Centiloid composite ROI

***Supplementary results***

*Main effect of CSF A$\beta_{42/40}$, age and APOE-ε4 on cortical amyloid deposition*

In both cohorts, Continuous CSF A$\beta_{42/40}$ was strongly and negatively associated to Aβ-PET uptake in widespread regions across the cortical mantle known to be target of early Aβ deposition (Chételat et al., 2013). Age was significantly related to a higher cortical Aβ deposition, even though the effect was much more prominent and widespread in ALFA, compared to ADNI. The effects of sex were marginal, and mostly indicating a higher deposition in woman compared to men in midline cortical areas including anterior and middle cingulate cortex, in both cohorts. There was no significant effect of *APOE*-ε4 on to Aβ-PET retention, however, removing CSF A$\beta_{42/40}$ from the model resulted in a significant main effect of the ε4 allele, indicating a higher Aβ deposition in carrier vs. non-carriers, across the cingulate cortex as well as in superior temporal areas (Fig. S2).

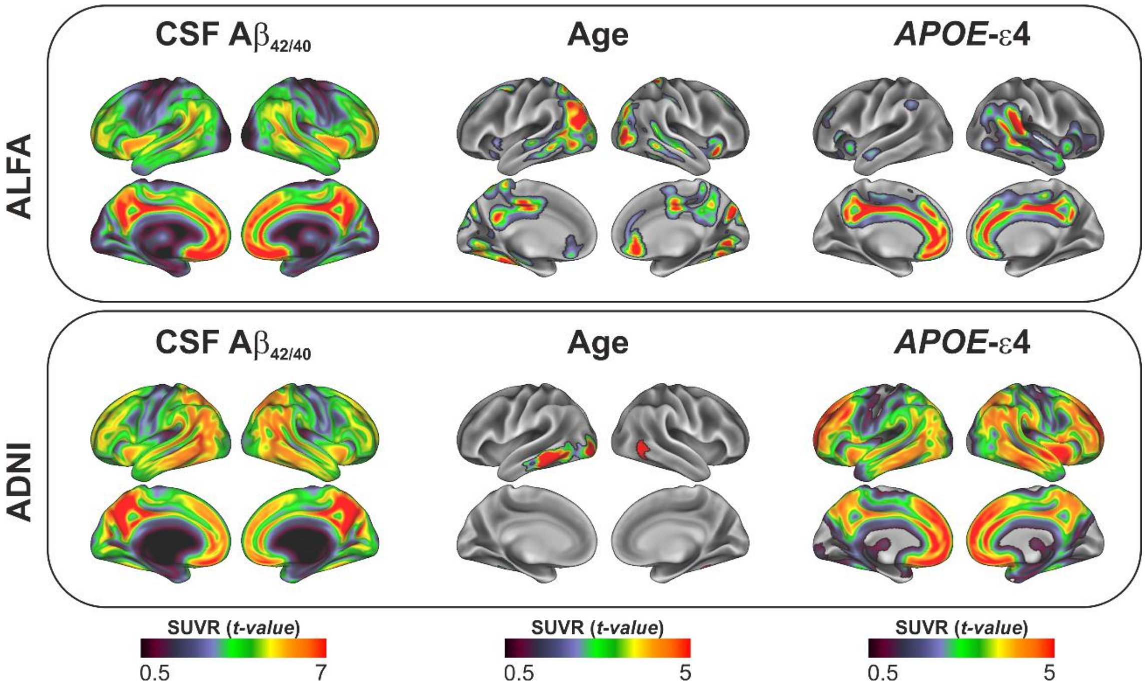


**Supplementary figure 2** – Main effects of CSF Aβ42/40, age and *APOE*-ε4 on Aβ PET tracer retention in both cohorts

**Supplementary Table 1** – List of Desikan-Killiany labels included in Braak stages and composite Centiloid ROIs

| | | **Desikan-Killiany labels** |
|---|---|---|
| **Centiloid** | | |
| | | Rostral middle frontal; Caudal middle frontal; Pars triangularis; Pars opercularis; Inferior parietal; Banks of superior temporal sulcus; middle temporal; Rostral anterior cingulate; Caudal anterior cingulate; Isthmus cingulate; Medial orbitofrontal; Putamen, Pallidum. |
| **Braak stage** | | |
| | I-II | Entorhinal cortex; Hippocampus |
| | III-IV | Fusiform; Lingual; Amygdala; Inferior temporal; Middle temporal; Temporal pole; Thalamus; Isthmus cingulate; Insula |
| | V-VI | Superior frontal; Paracentral; Precuneus; Cuneus; Pericalcarine; Rostral middle frontal; Caudal middle frontal; Precentral; Postcentral, Superior parietal; Lateral occipital; Transverse temporal; Banks of superior temporal sulcus; Superior temporal; Caudate; Putamen |
